# Automated Multilabel Diagnosis on Electrocardiographic Images and Signals

**DOI:** 10.1101/2021.09.22.21263926

**Authors:** Veer Sangha, Bobak J. Mortazavi, Adrian D. Haimovich, Antônio H. Ribeiro, Cynthia A. Brandt, Daniel L. Jacoby, Wade L. Schulz, Harlan M. Krumholz, Antonio Luiz P. Ribeiro, Rohan Khera

## Abstract

**Aims:** The application of artificial intelligence (AI) for automated diagnosis of electrocardiograms (ECGs) can improve access to high quality diagnostic care in remote settings but is limited by the reliance on signal-based data that are not routinely available. We sought to develop a multilabel automated diagnosis model for electrocardiographic images, more suitable for broader use.

**Methods and Results:** A total of 2,228,236 12-lead ECGs from 811 municipalities in Minas Gerais, Brazil were sampled into 90% (training):5%(validation):5%(held-out test), and were transformed to ECG images with varying lead locations and formats. We trained a convolutional neural network (CNN) using an EfficientNet-based architecture on ECG images to identify 6 physician-defined clinical labels spanning rhythm and conduction disorders, as well as a hidden label for gender. We trained another CNN for signal-based classification. The image-based model performed well on the held-out test set (average AUROC 0.99, AUPRC 0.68). This was replicated in a distinct test set from Brazil validated by at least two cardiologists (average AUROC 0.99, AUPRC 0.86) as well as an external validation set of 21,785 ECGs from Germany (average AUROC 0.97, AUPRC 0.73), with performance superior to signal-based models. Expert review of 120 out of 120 high confidence false positive predictions on the held-out and external validation sets were confirmed to be true positives with incorrect labels. The model learned generalizable features, confirmed using Gradient-weighted Class Activation Mapping (Grad-CAM).

**Conclusion:** We developed an externally validated model that extends the automated diagnosis of key rhythm and conduction disorders to printed ECGs as well as to the identification of hidden features, allowing the application of AI to ECGs captured across broad settings.

## BACKGROUND

An electrocardiogram (ECG) is an essential tool in the diagnosis and management of cardiovascular diseases, serving as an avenue for the identification of key pathophysiological signatures from the electrical activity of the heart. Currently, ECG data are collected as multichannel surface signal recordings of the cardiac electrical activity that are then transformed to images with printed waveforms. These images are then interpreted by trained clinicians, often precluding immediate diagnosis or the use of technology that can deliver deeper insights. While automated interpretation of ECGs promises to improve clinical workflow, particularly for key cardiovascular conditions, these tools are based on raw electrocardiographic signals^1,2^ rather than printed images.

Specifically, deep learning has been applied successfully to automate diagnosis based on signal data, performing comparably to trained clinicians for tasks such as detection of ECG abnormalities.^1-3^ However, a reliance on signal-based models poses a challenge in the real-world application of automated diagnosis, as ECGs are frequently printed and scanned as images. Thus, a major reorganization of operation is required to facilitate the application of models that focus on signals. Such technology is also inaccessible to paraclinical staff serving in remote settings, or to patients who increasingly have access to electrocardiographic images but lack ready access to experts for early diagnosis. Few tools have focused on automated diagnosis that allow for the incorporation of both ECG images and signals. Many existing models are trained and tested on data from a single source, with an inability to infer broad generalizability to different institutions and health settings. Finally, there has also been a preponderance of tools focusing on the diagnosis of single clinical entities,^4,5^ limiting clinical utility as ECGs can have multiple abnormalities simultaneously.

We created dual modality multilabel prediction algorithms that incorporate either ECG images or signals as inputs to predict the probability of various rhythm and conduction disorders using over 2 million ECGs from Brazil, independently validated in data from Germany.

## METHODS

### Data Source and Study Population

We used 12-lead ECGs collected by the Telehealth Network of Minas Gerais (TNMG) and described previously in Ribeiro, et. al. (2020).^3^ The data were assembled as a part of the Clinical Outcomes in Digital electrocardiography (CODE) study.^6^ They include deidentified signal data captured between 2010 and 2017 from 811 out of the 853 municipalities in the state of Minas Gerais, Brazil. The ECGs obtained were collected using one of two models of tele-electrocardiograph machines, the TEB ECGPC, manufactured by Tecnologia Electronica Brasileira, or the ErgoPC 13, manufactured by Micromed Biotecnologia. These ECGs were recorded as standard 12-lead recordings sampled at frequencies ranging from 300 to 600 hz for 7 to 10 seconds. Additionally, information on patient demographics and 6 clinical labels were available. Further, to augment the evaluation of models built on the primary CODE study dataset, where clinical annotations were derived from routine clinical care, a secondary cardiologist-validated annotation test dataset was used. This consisted of additional ECGs obtained from the TNMG network between April and September 2018. These ECGs were rigorously validated by 2-to-3 independent cardiologists based on criteria from the American Heart Association.^7^ Finally, an ECG dataset from Germany was obtained for external validation (described in section on external validation).

### Data Preprocessing

We resampled all ECGs to a 300 Hz sampling rate with 5 second simultaneous recordings across leads to obtain uniform length inputs. Each sample started 300ms before the second QRS peak detected by the XQRS algorithm in lead I of the original recording.^8^ ECGs that did not have any peaks detected (94,277, or 4.1%) were discarded, as inspection of a sample of 50 ECGs after further preprocessing did not detect any true cardiac activity.

For signal data, each ECG was processed as a matrix of 12 leads of 1500 sequential data points, representing a 5 second acquisition at a 300 Hz sampling frequency. The 5 second samples were preprocessed with a one second median filter applied across each lead and subtracted from the original waveform to remove baseline drift. Next, we employed a peak annotation technique for each ECG to include ECG morphology along with waveform data as an input to the signal model. For all ECGs, we found the median and standard deviation of all RR, PR, QRS, and ST intervals for each lead using NeuroKit2, a Python toolkit for signal processing.^9^ The median lengths of each interval across leads were included as additional inputs into the signal model (**Figure 1**).

**Figure 1:**
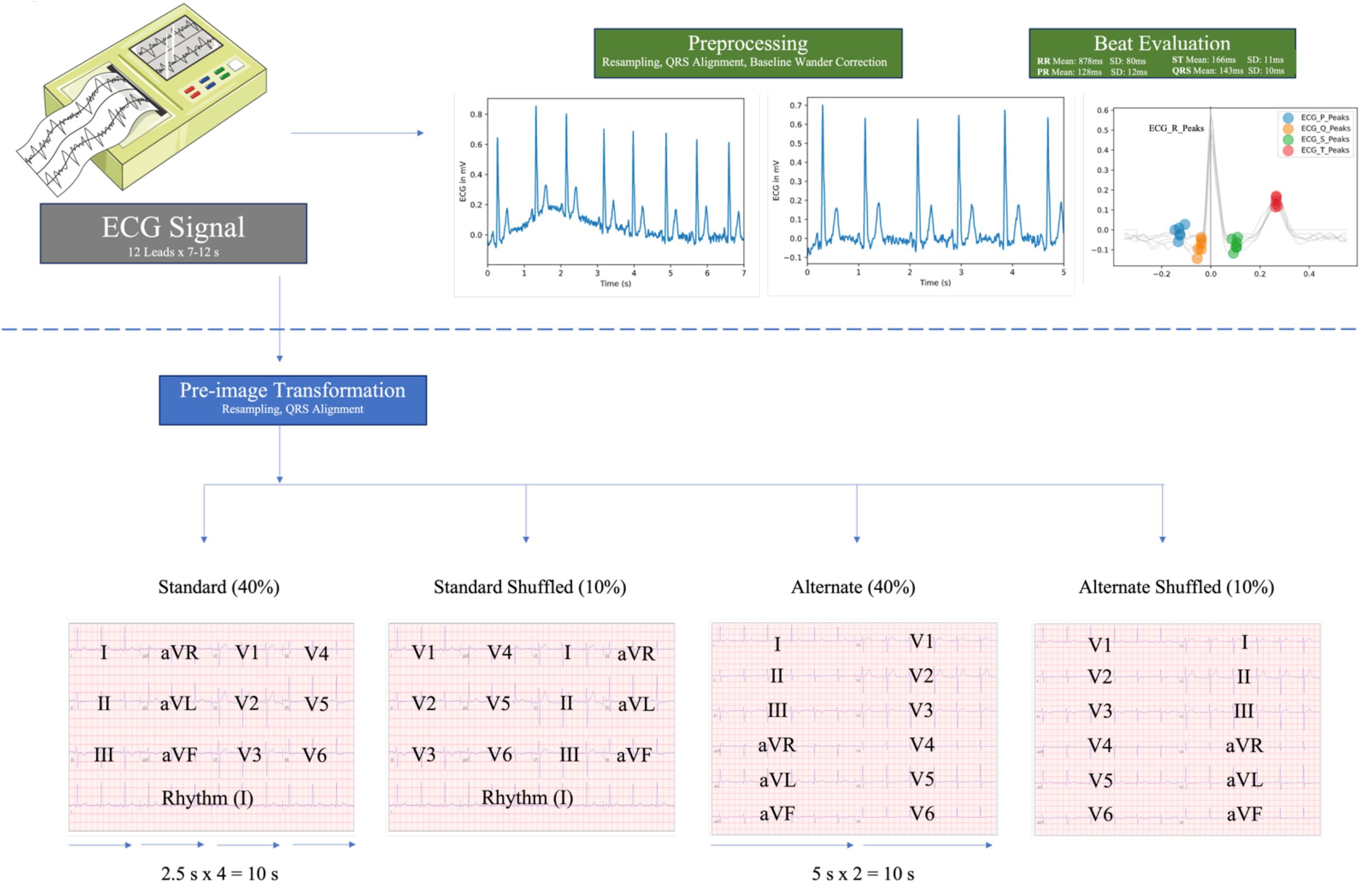
Study Outline

The ECGs were transformed to the corresponding printed images using the python library ecg-plot.^10^ To ensure that like a clinician the model learned about lead-specific information based on the label of the lead, rather than just the location of leads on a single ECG format, we trained on four formats of images (**Figure 1**). Standard printed ECGs in the United States typically consist of four 2.5 second columns presented sequentially on the page, with each column containing three leads of 2.5 second intervals of a continuous 10 second record. A 10-second rhythm strip was generated using concatenation of lead I signal on the last and first QRS of the same 5 second signal.^7^ The second plotting scheme we chose consisted of two columns each containing six 5-second recordings, with one column containing simultaneous limb leads and the other simultaneous precordial leads. We treated this scheme as our alternate image form (**Figure 1**).

The third and fourth formats were representations of the standard and alternate formats with shuffled lead locations. In the shuffled standard format precordial leads were presented in the first two columns and limb leads in the third and fourth. In shuffled alternative images precordial leads were presented in the first column and limb leads in the second (**Figure 1**).

### Study Outcomes

Each ECG was annotated for 6 physician-defined clinical labels spanning rhythm and conduction disorders (atrial fibrillation (AF), right bundle branch block (RBBB), left bundle branch block (LBBB), 1^st^ degree AV block (1dAVB), sinus tachycardia (ST), and sinus bradycardia (SB)). Diagnoses for ECGs were obtained by a combination of the automated diagnosis provided by the Glasgow ECG analysis software and the natural language processing (NLP) extracted diagnosis from a cardiologist report of the ECGs. To detect whether our image and signal-based modeling approaches could detect hidden or higher order features that are not discernable from the ECG by human readers, we defined patient sex as the seventh label given its consistent availability across the study population and prior literature describing its detection from signal-based electrocardiographic deep learning models.^11^

### Experimental Design

All ECGs were randomly subset into training, validation, and test sets (90%-5%-5%). Given the low prevalence of all clinical labels, the data splits were stratified by clinical labels, so cases of each of the 6 labelled clinical disorders were split proportionally among the sets, as was gender. To ensure that model learning was not affected by the low frequency of certain labels, custom loss functions based on the effective number of samples class sampling scheme were used for both image and signal models, with weighting based on the number of samples for each class (**eTable 2**).^12^

### Image Model Overview

For image data, we built a convolutional neural network model based on the EfficientNet set of architectures.^13^ To balance the complexity and accuracy of a model with the computational resources required for training, we chose the B3 architecture, which required PNG images to be sampled at 300 × 300 square pixels. The model includes 384 layers, has over 10 million trainable parameters, and is composed of 7 blocks (**eFigure 1**). Additional dropout layers were added to prevent overfitting, and the output of the EfficientNet-B3 model was fed into a fully connected layer with sigmoid activation to predict the probability of target labels. The model was trained with an Adagrad optimizer and learning rate 5 ⨯ 10^−3^ for 7 epochs with a minibatch size of 64. The optimizer and learning rate were chosen after hyperparameter optimization. We *a priori* chose 30 epochs for training models with built in early stopping based on validation loss not improving after 3 consecutive epochs. Model weights were initialized as the pretrained EfficientNetB3 weights on the ImageNet dataset to take advantage of any possible transfer learning.

We trained and validated our primary image model with a 40%-40%-10%-10% split of standard, alternate, standard shuffled, and alternate shuffled images among the training and validation sets, stratified in the same manner described above (see schematic in **Figure 1**). This was done to prevent overfitting to certain locations on the ECG image, and to create a more versatile algorithm. In sensitivity analyses, we trained and validated on a 50%-50% split of standard and alternate images to evaluate whether the introduction of shuffling of the leads had improved model performance.

### Signal Model Overview

For signal data, we developed a custom convolutional neural network (CNN) model that combined inception blocks, convolutional, and fully connected layers, similarly to Raghunath et al.^4^ Our model had two inputs, the 1500×12 waveform data, and the 8×1 array of derived elements from the ECG including various standard intervals.

The waveform data input was passed through 7 branches, each one containing an initial 1D-convolutional layer followed by 5 inception blocks and another 1D-convolutional layer (**eFigure 2**). We used inception blocks to capture information of various kernel sizes and therefore differing assumptions of the locality, or spatial connectivity of ECG data. For example, smaller kernels performed more localized learning, on individual waves or parts of waves, while larger ones combined data across sections of multiple waves.

The signal in each of these branches was then flattened, passed through two fully connected layers, and concatenated with data from the other branches. These final waveform signal data were passed through a fully connected layer and then concatenated with the 8×1 peak morphology input. This information was passed through two more fully connected layers, the second of which had sigmoid activation to predict the probability of target labels. Further details of the signal models are included in **eMethods** (**Online Supplement)**.

In sensitivity analyses, we trained a signal model with the exact same architecture as the one described, but without peak morphology inputs to test the ability of a convolutional network to perform these operations internally and learn the same patterns.

### Internal and External validation

In addition to the held-out test set, we validated the algorithm in an expert validated test of 827 ECGs from distinct patients in Minas Gerais, Brazil during a different time window (April to September 2018). The labels on these ECGs were validated by 2 cardiologists, with disagreements resolved by a 3^rd^ cardiologist, based on AHA criteria.

We also pursued external validation to assess the ability to generalize to novel data sources.^14^ In addition to the held-out test set, model performance was evaluated on the Germany-based external validation dataset, PTB-XL, whose data have been previously described.^15^ Briefly, the dataset has 21,837 recordings from 18,885 patients. ECGs were collected with devices from Schiller AG between 1989 and 1996 and are available as 10 second recordings sampled at 500 hz for 10 seconds. Each record has labels for diagnostic, form, and rhythm statements. Data were transformed in the same manner as the data from Brazil, and the same labels were extracted to assess model performance.

### Model Interpretability

We used Gradient-weighted Class Activation Mapping (Grad-CAM) to highlight the regions in an image predicting a given label.^16^ This provides interpretability of the model’s predictions and allows for the evaluation of whether model-assigned labels are based on clinically relevant information or on heuristics based on spurious data features.^17^ To deploy Grad-CAM in our models, we calculated the gradients of the final stack of filters in the network for each prediction class of interest. These gradients assigned the importance of a given pixel to the prediction of the label. Then, we created filter importance weights by performing a global average pooling of the gradients in each filter, emphasizing filters whose gradient suggested they contributed to the prediction of the class of interest. Finally, each filter in our final convolutional layer was multiplied by its importance weight and combined across filters to generate a Grad-CAM heatmap. We overlayed the heatmaps on the original ECG images.

We used two approaches to assess model interpretability. First, we examined individual ECGs, obtaining, and overlaying the Grad-CAM heatmaps for both the standard and alternate images. Second, we averaged class activation maps for the 25 most confident predictions for a condition of interest for any given model. We simply ranked images by the output of our model for that image, computed a simple mean across the heatmaps for the 25 images and overlayed this average heatmap over a representative ECG to understand it in context.

We chose LBBB and RBBB to illustrate the interpretability as these labels have lead-specific information that is used to make the clinical diagnosis. This contrasts with the other labels (AF, ST, SB, 1dAVB) where the information on these rhythm disorders can be deduced from any of the ECG leads, limiting assessment of lead-specific learning.

### Statistical Analysis

Model performance was evaluated in the held-out test set, cardiologist-validated test set, and the external validation set. We assessed area under the receiver operating characteristic (AUROC) curve, which represents model discrimination, with values ranging from 0.50 to 1.00, representing random classification and perfect discrimination of labels, respectively. In addition, we assessed the area under the precision recall curve (AUPRC), and the sensitivity, specificity, positive predictive value, negative predictive value, and F1 score of the model for each label. For the threshold dependent measures, the threshold that maximized the F1 score was selected, representing a strategy that optimized both precision and recall. Class weighted mean metrics across clinical labels were calculated to evaluate the performance of a model at a dataset-level by taking the weighted average of metrics on the labels in that dataset (e.g. held-out test set, cardiologist-validated internal test set, external validation set), weighed by the counts of labels in that dataset. Models were compared by computing p values and 0.95 confidence intervals for AUROC using DeLong’s Method.^18^ We used confusion matrices to illustrate the discordance between a model’s predictions and the diagnoses that came with our datasets. These were constructed among ECGs with single clinical labels.

### Manual Review of False Positive Predictions

Two cardiologists independently reviewed sample of 10 false positives for each clinical label with the highest probability of a given label (n = 120) to verify the accuracy of the labels and qualitatively assess the potential ECG features that may have prompted a false positive result in both the held-out test set and the external validation data.

All analyses were performed using Python 3.9. The study was reviewed by the Yale Institutional Review Board, which waived informed consent as the study uses deidentified data.

## RESULTS

### Study Population

There were 2,228,236 ECGs from 1,506,112 patients acquired between 2010-2017. The median age of the patients at the time of the ECG recording was 54 years (IQR 41, 67) and 60.3% of the ECGs were obtained among women.

Of these ECGs, 39,661 (1.8%) had a label for AF, 61,551 (2.8%) for RBBB, 34,677 (1.6%) for LBBB, 34,446 (1.5%) for 1dAVb, 35,441 (1.6%) for SB, and 48,296 (2.2%) for ST (**eTable 1)**. Of the 231,704 ECGs with at least one of the six detected rhythm disorders, 210,496 had exactly one, and 21,208 had more than one label.

### Performance of Image-based Multilabel Classification

The AUROCs for clinical labels on the held-out test set ranged from 0.97 for 1dAVb to 1.00 (0.996) for LBBB (**Table 1**). AUROC was greater than 0.99 for RBBB, LBBB, SB, AF, and ST, but was lower for 1dAVB. The AUPRCs for clinical labels ranged from 0.37 for 1dAVb to 0.81 for LBBB. AUPRC was greater than 0.69 for RBBB, LBBB, AF, and ST, with lower values for SB and 1dAVB (**Table 1**). At cutoffs that ensured maximum F1 value, specificity was above 0.88 for all clinical labels, and sensitivity was above 0.75 for RBBB, LBBB, SB, AF, and ST, but was lower for 1dAVb (0.48) (**Table 1**). For the higher order label of gender, AUROC was 0.91, AUPRC 0.87, and specificity and sensitivity 0.83 and 0.83 respectively.

**Table 1:** Performance of Image and Signal Based Models on Held-out test set and Cardiologist Validated Test Set.

| Data Source | Model | Label | Accuracy | PPV | NPV | Specificity | Sensitivity | AUROC | F1 | AUPRC |
| --- | --- | --- | --- | --- | --- | --- | --- | --- | --- | --- |
| **Held-out Test Set** | **Image** | Male | 0.831 | 0.766 | 0.879 | 0.834 | 0.826 | 0.910 (0.908 - 0.911) | 0.795 | 0.873 |
|  |  | 1dAVb | 0.98 | 0.39 | 0.992 | 0.988 | 0.486 | 0.968 (0.965 - 0.971) | 0.433 | 0.372 |
|  |  | RBBB | 0.987 | 0.725 | 0.996 | 0.991 | 0.856 | 0.993 (0.992 - 0.994) | 0.785 | 0.786 |
|  |  | LBBB | 0.993 | 0.734 | 0.997 | 0.995 | 0.832 | 0.996 (0.996 - 0.997) | 0.78 | 0.805 |
|  |  | SB | 0.985 | 0.534 | 0.996 | 0.989 | 0.778 | 0.989 (0.988 - 0.991) | 0.633 | 0.581 |
|  |  | AF | 0.989 | 0.668 | 0.996 | 0.993 | 0.755 | 0.988 (0.986 - 0.990) | 0.709 | 0.737 |
|  |  | ST | 0.986 | 0.624 | 0.997 | 0.989 | 0.848 | 0.993 (0.993 - 0.994) | 0.719 | 0.693 |
|  |  | Weighted Mean | 0.987 | 0.625 | 0.996 | 0.991 | 0.774 | 0.989 (0.987 - 0.990) | 0.691 | 0.678 |
|  | **Signal** | Male | 0.704 | 0.594 | 0.829 | 0.641 | 0.8 | 0.803 (0.801 - 0.806) | 0.682 | 0.737 |
|  |  | 1dAVb | 0.968 | 0.214 | 0.991 | 0.976 | 0.411 | 0.934 (0.930 - 0.938) | 0.282 | 0.197 |
|  |  | RBBB | 0.987 | 0.733 | 0.995 | 0.992 | 0.827 | 0.987 (0.986 - 0.989) | 0.777 | 0.745 |
|  |  | LBBB | 0.992 | 0.697 | 0.997 | 0.994 | 0.821 | 0.994 (0.993 - 0.995) | 0.754 | 0.754 |
|  |  | SB | 0.986 | 0.561 | 0.996 | 0.99 | 0.767 | 0.990 (0.990 - 0.991) | 0.648 | 0.589 |
|  |  | AF | 0.984 | 0.535 | 0.994 | 0.99 | 0.643 | 0.979 (0.976 - 0.982) | 0.584 | 0.567 |
|  |  | ST | 0.986 | 0.639 | 0.997 | 0.989 | 0.854 | 0.992 (0.991 - 0.994) | 0.731 | 0.708 |
|  |  | Weighted Mean | 0.984 | 0.584 | 0.995 | 0.989 | 0.738 | 0.981 (0.979 - 0.983) | 0.649 | 0.615 |
| **Cardiologist-Validated Test Set** | **Image** | Male | 0.775 | 0.669 | 0.874 | 0.739 | 0.832 | 0.845 (0.818 - 0.872) | 0.742 | 0.775 |
|  |  | 1dAVb | 0.985 | 0.808 | 0.991 | 0.994 | 0.75 | 0.989 (0.981 - 0.997) | 0.778 | 0.744 |
|  |  | RBBB | 0.982 | 0.721 | 0.996 | 0.985 | 0.912 | 0.979 (0.956 - 1.000) | 0.805 | 0.832 |
|  |  | LBBB | 0.996 | 0.966 | 0.997 | 0.999 | 0.933 | 1.000 (0.999 - 1.000) | 0.949 | 0.989 |
|  |  | SB | 0.992 | 0.737 | 0.998 | 0.994 | 0.875 | 0.995 (0.991 - 0.999) | 0.8 | 0.756 |
|  |  | AF | 0.992 | 0.75 | 0.995 | 0.996 | 0.692 | 0.996 (0.992 - 1.000) | 0.72 | 0.786 |
|  |  | ST | 0.987 | 0.81 | 0.996 | 0.99 | 0.919 | 0.997 (0.994 - 0.999) | 0.861 | 0.927 |
|  |  | Weighted Mean | 0.988 | 0.808 | 0.995 | 0.992 | 0.867 | 0.992 (0.984 - 0.999) | 0.833 | 0.857 |
|  | **Signal** | Male | 0.712 | 0.594 | 0.847 | 0.646 | 0.816 | 0.795 (0.764 - 0.825) | 0.688 | 0.703 |
|  |  | 1dAVb | 0.97 | 0.538 | 0.991 | 0.977 | 0.75 | 0.970 (0.947 - 0.993) | 0.627 | 0.607 |
|  |  | RBBB | 0.99 | 0.842 | 0.997 | 0.992 | 0.941 | 0.994 (0.989 - 1.000) | 0.889 | 0.881 |
|  |  | LBBB | 0.996 | 0.966 | 0.997 | 0.999 | 0.933 | 0.997 (0.993 - 1.000) | 0.949 | 0.967 |
|  |  | SB | 0.989 | 0.667 | 0.998 | 0.991 | 0.875 | 0.996 (0.992 - 1.000) | 0.757 | 0.814 |
|  |  | AF | 0.99 | 0.667 | 0.996 | 0.994 | 0.769 | 0.986 (0.966 - 1.000) | 0.714 | 0.746 |
|  |  | ST | 0.994 | 0.944 | 0.996 | 0.997 | 0.919 | 0.998 (0.996 - 1.000) | 0.932 | 0.952 |
|  |  | Weighted<br>Mean | 0.988 | 0.803 | 0.996 | 0.992 | 0.88 | 0.991 (0.982 - 0.999) | 0.836 | 0.848 |

Performance on standard and alternate format images was comparable. The class mean weighted AUROC across clinical labels on the held-out test set images in both standard and alternate format was 0.99 (**eTable 3**). Class mean weighted AUPRC across clinical labels was 0.68 on images for the standard format, and 0.69 for images in the alternate format. For the higher order label of gender, AUROC was 0.91 for both standard and alternate format images, and AUPRC was 0.87 for standard format images and 0.88 for alternate format images.

### Performance of Signal-based Multilabel Classification

The AUROCs for clinical labels on the held-out test set ranged from 0.93 for 1dAVb to 0.99 for LBBB. AUROC was greater than 0.98 for RBBB, LBBB, SB, AF, and ST, but was lower for 1dAVB (**Table 1**). The AUPRCs for clinical labels ranged from 0.20 for 1dAVb to 0.75 for LBBB. AUPRC was greater than 0.70 for RBBB, LBBB, and ST, and greater than 0.55 for SB and AF. At cutoffs that corresponded to the highest F1 value, specificity was above 0.98 for all clinical labels, and sensitivity was above 0.75 for RBBB, LBBB, SB, AF, and ST, but was lower for 1dAVb (0.41). For the higher order label of gender, AUROC was 0.80, AUPRC 0.74, and specificity and sensitivity 0.64 and 0.80 respectively.

### Internal and External Validation

The class weighted mean AUROC across clinical labels on the cardiologist-validated internal test set was higher than the held-out test. For the image-based model, it was 0.99 (0.95 CI 0.98 – 1.00), and for the signal-based model it was 0.99 (0.95 CI 0.98 – 1.00) (**Table 1**). Class weighted mean AUPRCs were also higher on the cardiologist-validated test set, 0.86 for the image-based model and 0.85 for the signal-based model. Performance on the higher order label of gender was slightly lower for the image-based model, with an AUROC of 0.85 (0.95 CI 0.92 – 0.87) and AUPRC of 0.78 but was comparable for the signal-based model (AUROC 0.80 (0.95 CI 0.76 0 0.83) and AUPRC 0.70).

In the external validation set, PTB-XL, model performance was comparable to the held-out test set. For the image-based model, class weighted mean AUROC was 0.97 (0.95 CI 0.97 – 0.98) and for the signal-based model it was 0.96 (0.95 CI 0.95 – 0.96) (**Table 2**). Class weighted mean AUPRCs were also comparable, 0.73 for the image-based model and 0.65 for the signal-based model. Performance on the higher order label gender was also comparable across datasets. For the image-based model, AUROC was 0.87 (0.95 CI 0.85 – 0.87) and AUPRC was 0.88 on PTB-XL, and for the signal-based model, AUROC was 0.74 (0.95 CI 0.73 – 0.75), and AUPRC was 0.75.

**Table 2:** Performance of Image and Signal Based Models on the external validation set PTB-XL

| Model | Label | Accuracy | PPV | NPV | Specificity | Sensitivity | AUROC | F1 | AUPRC |
| --- | --- | --- | --- | --- | --- | --- | --- | --- | --- |
| **Image** | Male | 0.774 | 0.736 | 0.837 | 0.655 | 0.883 | 0.869 (0.845 - 0.874) | 0.802 | 0.877 |
|  | 1dAVb | 0.941 | 0.316 | 0.982 | 0.956 | 0.532 | 0.920 (0.912 - 0.927) | 0.396 | 0.331 |
|  | RBBB | 0.987 | 0.694 | 0.996 | 0.99 | 0.85 | 0.994 (0.992 - 0.995) | 0.764 | 0.77 |
|  | LBBB | 0.993 | 0.829 | 0.997 | 0.996 | 0.875 | 0.996 (0.994 - 0.997) | 0.852 | 0.907 |
|  | SB | 0.981 | 0.687 | 0.99 | 0.991 | 0.666 | 0.963 (0.956 - 0.969) | 0.676 | 0.648 |
|  | AF | 0.973 | 0.788 | 0.988 | 0.983 | 0.833 | 0.985 (0.982 - 0.988) | 0.81 | 0.867 |
|  | ST | 0.983 | 0.761 | 0.992 | 0.99 | 0.794 | 0.990 (0.988 - 0.992) | 0.777 | 0.806 |
|  | Weighted Mean | 0.974 | 0.686 | 0.99 | 0.983 | 0.761 | 0.974 (0.971 - 0.978) | 0.718 | 0.733 |
| **Signal** | Male | 0.651 | 0.615 | 0.759 | 0.398 | 0.883 | 0.740 (0.733 - 0.746) | 0.725 | 0.748 |
|  | 1dAVb | 0.919 | 0.19 | 0.976 | 0.939 | 0.377 | 0.862 (0.852 - 0.872) | 0.253 | 0.177 |
|  | RBBB | 0.985 | 0.66 | 0.996 | 0.989 | 0.847 | 0.986 (0.981 - 0.992) | 0.742 | 0.729 |
|  | LBBB | 0.991 | 0.819 | 0.996 | 0.996 | 0.824 | 0.995 (0.993 - 0.997) | 0.821 | 0.87 |
|  | SB | 0.98 | 0.659 | 0.99 | 0.989 | 0.68 | 0.955 (0.947 - 0.963) | 0.669 | 0.642 |
|  | AF | 0.957 | 0.676 | 0.981 | 0.974 | 0.742 | 0.972 (0.969 - 0.975) | 0.707 | 0.729 |
|  | ST | 0.982 | 0.745 | 0.991 | 0.99 | 0.779 | 0.987 (0.983 - 0.990) | 0.762 | 0.794 |
|  | Weighted Mean | 0.965 | 0.619 | 0.986 | 0.977 | 0.701 | 0.958 (0.953 - 0.963) | 0.653 | 0.653 |

### Comparison of Performance of Image and Signal Models

The image and signal models performed comparably for clinical labels on both datasets, with high correlation between prediction across labels. For the clinically important diagnosis of AF, the image-based model had AUROCs of 0.99 (0.95 CI 0.99 – 0.99) on the held-out test set, 1.00 (0.95 CI 0.99 – 1.00) on the cardiologist-validated internal test, and 0.99 (0.95 CI 0.98 – 0.99) on PTB-XL, while the signal-based model had AUROCs of 0.98 (0.95 CI 0.98 – 0.98), 0.99 (0.95 CI 0.97 – 1.00) and 0.97 (0.95 CI 0.97 – 0.98) respectively. The class weighted mean AUROC across clinical labels was also comparable; 0.99 (0.95 CI 0.99 – 9.99) on the held-out test set, 0.99 (0.95 CI 0.98 – 1.00) on the cardiologist-validated internal test set, and 0.98 (0.95 CI 0.97 – 0.98) on PTB-XL for the image-based model, and 0.98 (0.95 CI 0.98 – 0.98), 0.99 (0.95 CI 0.98 – 1.00), and 0.96 (0.95 CI 0.95 – 0.96) for the signal-based model (**Table 1, 2**). For the higher order label of gender, the image-based model outperformed the signal-based one, with AUROC of 0.91, 0.85 and 0.87 on the held-out test, cardiologist-validated test, and PTB-XL, respectively, compared with 0.80, 0.80, and 0.74 for the signal-based model (p < .001 for difference on held-out test, PTB-XL). The high discrimination across labels and in all three datasets for both image and signal-based models was noted in ROC curves (**Figure 2, 3**).

**Figure 2:**
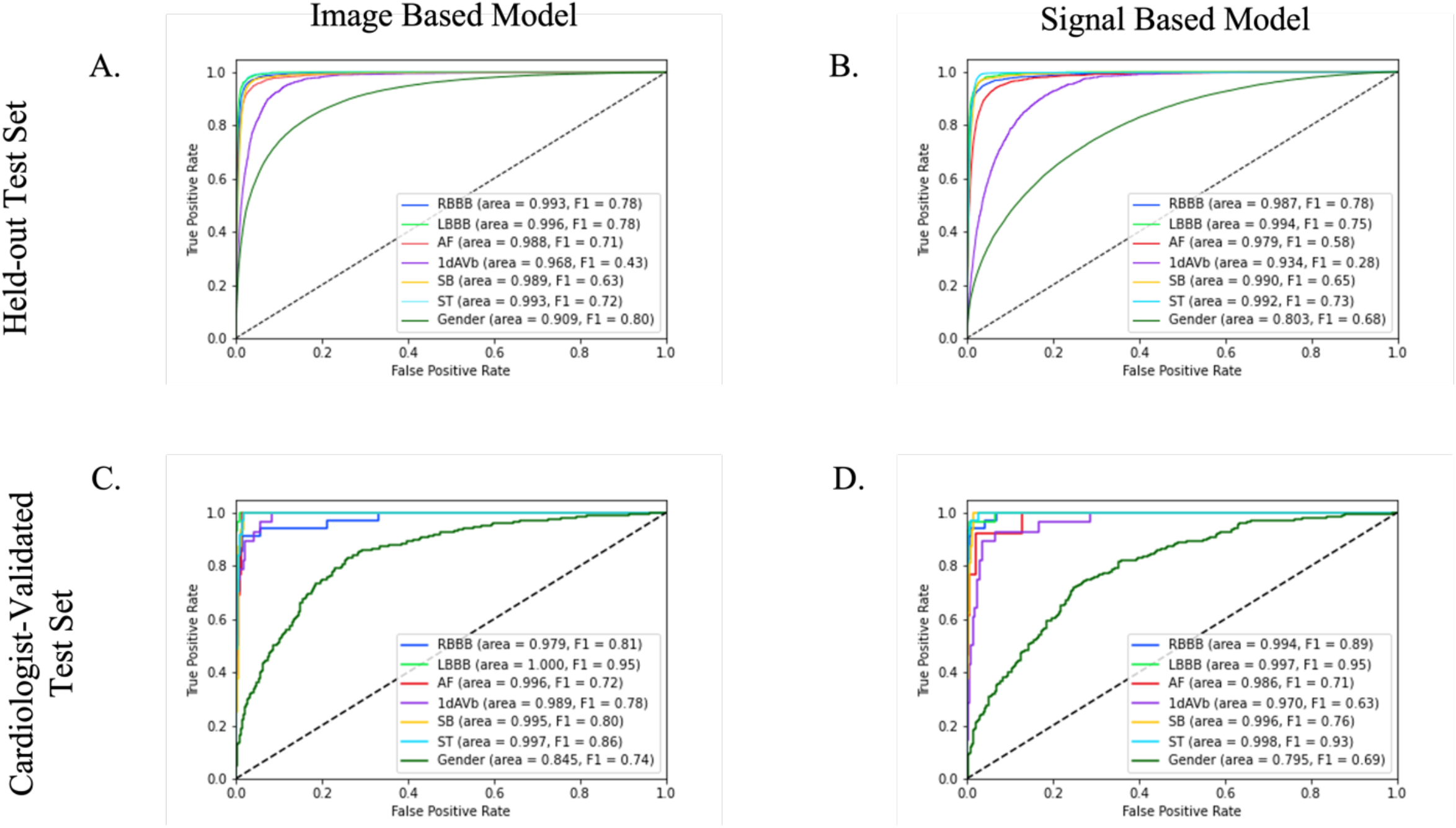
ROC Curves for Image and Signal based models on Held-out Test set and Cardiologist Validated Test set

**Figure 3:**
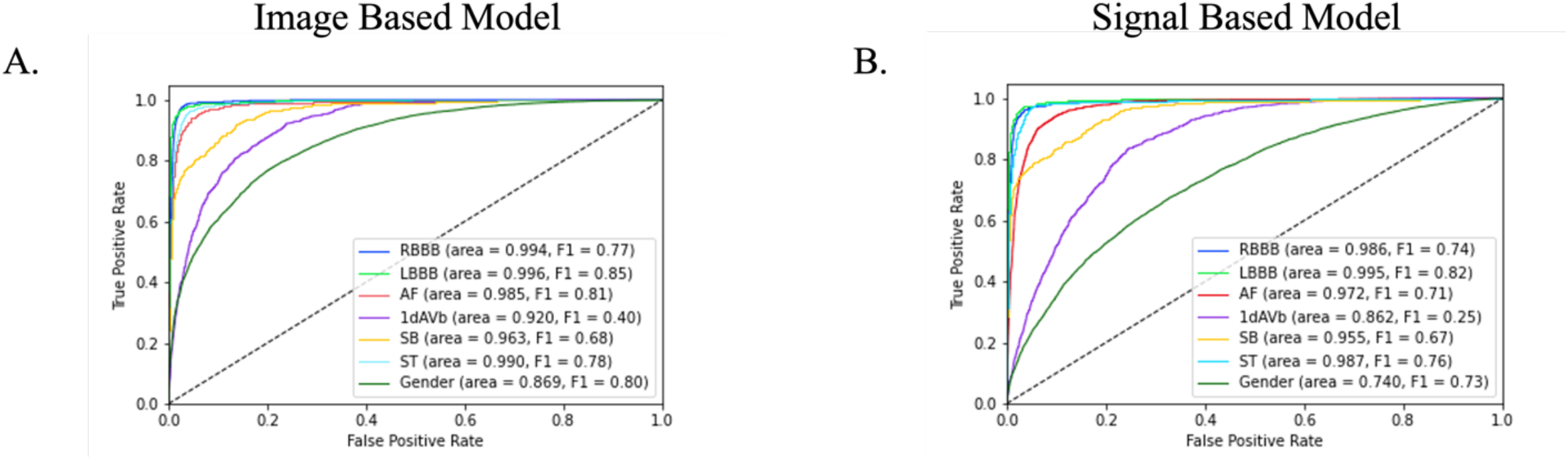
ROC Curves for Image and Signal based models on PTB-XL

The label-level performance of image and signal based models was also consistent, with the highest AUROC and AUPRC scores on the same clinical labels, LBBB and RBBB, (AUROC of 1.00 (0.996) and 0.99 for image-based, and 0.99 and 0.99 for signal based, and AUPRC of 0.81 and 0.79 for image-based and 0.75 and 0.75 for signal-based on the held-out test set) and lowest scores on the same class, 1dAVb (AUROC of 0.97 for image-based, and 0.93 for signal based, and AUPRC of 0.37 for image-based, and 0.20 for signal-based on the held-out test set). Confusion matrices showed that among ECGs with only one clinical label, predictions of LBBB, RBBB, and ST were the most accurate for both image and signal based models (above 83% for all three for the image-based model, and above 81% for the signal-based one) **(Figure 4, eFigure 3)**. These findings were consistent in the cardiologist-validated set and PTB-XL.

**Figure 4:**
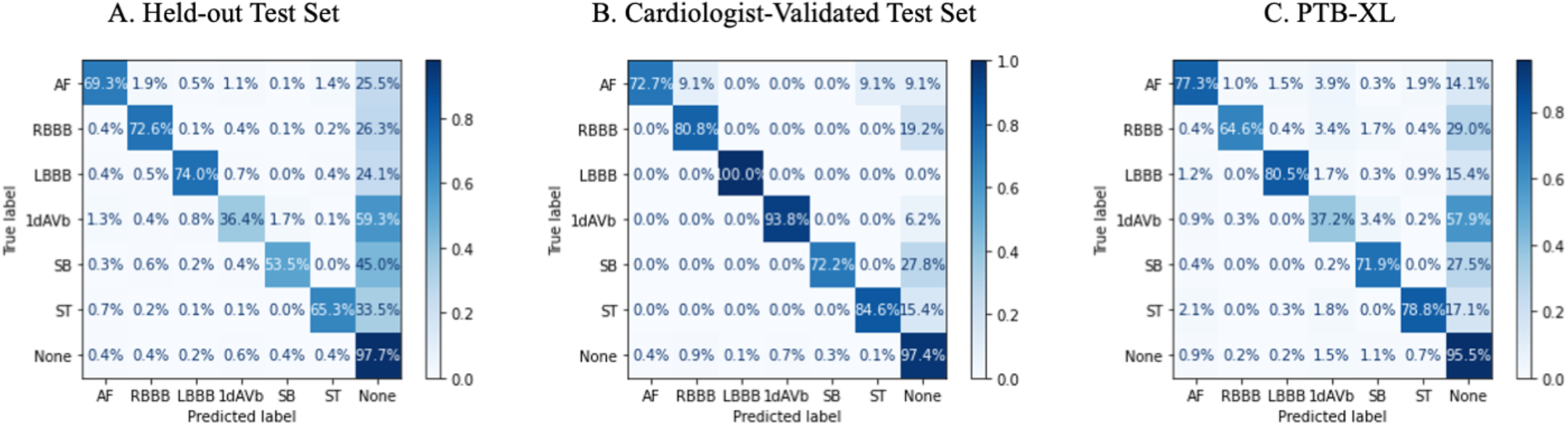
Confusion Matrices for Image Model Predictions

### Manual Review for Misclassified ECGs

The most common errors across algorithms were type 1, or false positives. We took 120 false positives from our image-based model with the highest probability for each clinical label in the held-out test set and external validation data, PTB-XL (10 for each label in each dataset). Expert review by cardiologists confirmed that all 120 were accurately classified by the model and had incorrect labels i.e., that these were true positive results (ECGs available at https://github.com/CarDS-Yale/ECG-DualNet).

### Prediction Interpretability with Grad-CAM

The Grad-CAMs identified sections of the ECG that were most important for the label classification. **Figure 5** shows the average class activation heatmaps for RBBB and LBBB predictions on standard and alternate form images, highlighting ECG regions that were important for the diagnosis of each rhythm across many predictions. The region of the ECG corresponding to lead V4 was the most important for prediction of RBBB across both the standard and alternate images, and the region corresponding to lead V2 was also equally important for prediction of RBBB across standard image. On the other hand, while regions corresponding to lead I and lead V3 were most important for the prediction of LBBB across standard images, regions corresponding to lead V4 and aVR were most important for LBBB predictions across alternate images. **eFigure 4** shows Grad-CAMs for individual representative examples of model prediction of RBBB, LBBB, SB, and ST on standard images.

**Figure 5:**
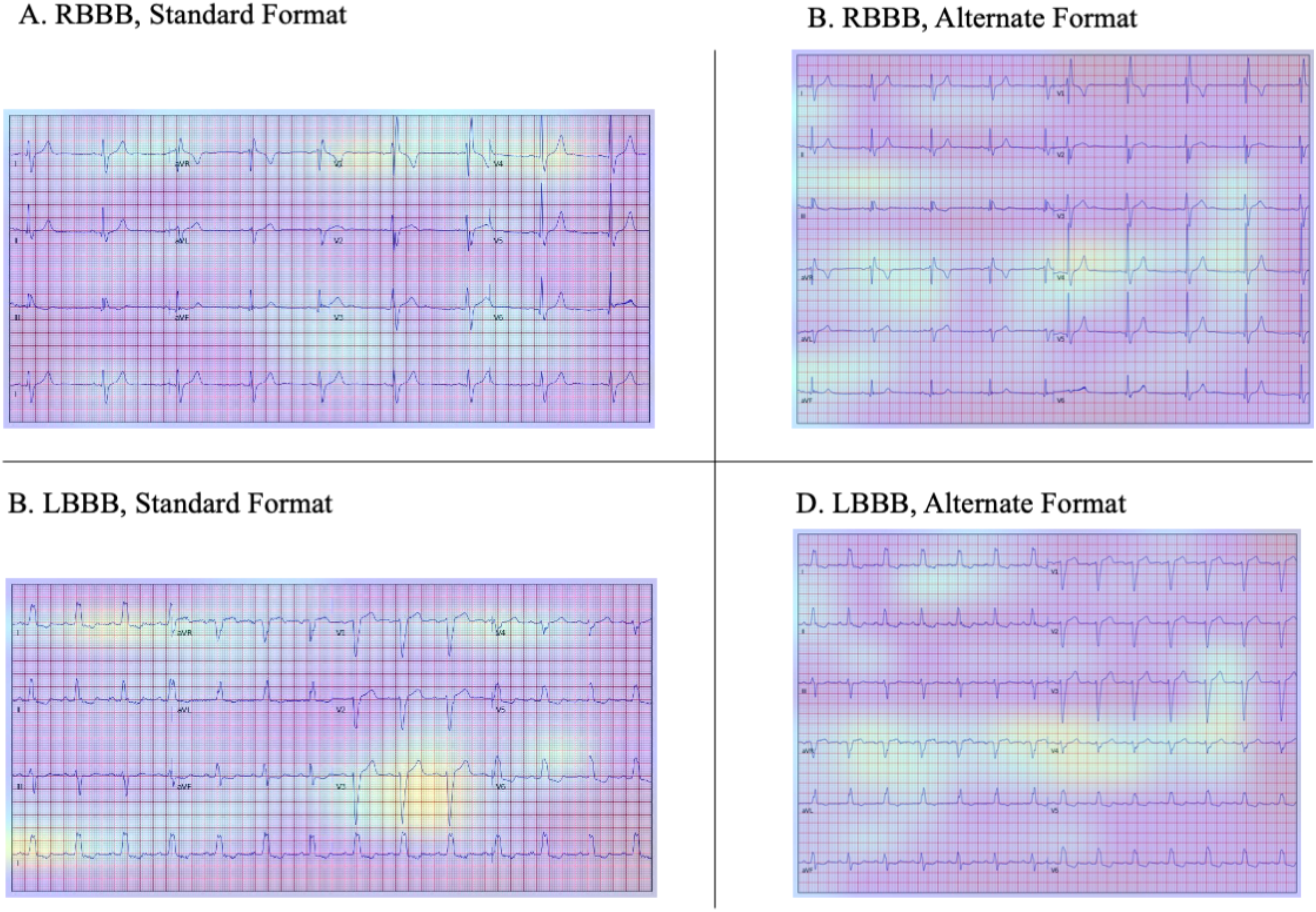
Average of the Grad-CAMs for the 25 most confident predictions of RBBB and LBBB on standard and alternate format images

### Sensitivity analyses

For our image-based models – the one trained with shuffled data, and the one trained with only traditional printed inputs – performed comparably on both standard and alternate form printed images (**eTable 3 and 4**). Our signal model without peak morphology information also performed comparably to the signal model with them **(eTable 5 and 6**).

## DISCUSSION

We have developed an externally validated multilabel automated diagnosis algorithm that accurately identifies rhythm and conduction disorders from either ECG images or raw electrocardiographic signals. The algorithm demonstrates high discrimination and generalizes across two international data sources, which are acquired on different equipment and temporally separated by over 2 decades. The models are also invariant to the layout of the ECG images, with interpretable recognition of leads of interest and abnormalities by the image-based algorithms. The models demonstrate that ECG images, like signals, can be used to define hidden features, such as gender, which has thus far exclusively been done with signal-based models. Our approach has the potential to broaden the application of artificial intelligence to electrocardiographic data across storage strategies.

Image-based models represent an important advance in automated diagnosis from ECGs as they allow applications to data sources for which raw signals may not be available. This represents most healthcare settings that have not been optimized for storing and processing signal data in real-time and that rely on printed or scanned ECG images. In addition, in contrast to ECG-based models that have been developed in single health systems,^2,11,19^ our models have broad external validity, performing equivalently in regionally and temporally distinct validation data. The image-based models also demonstrate comparable performance to signal-based models in published reports^2,20^ despite both the substantial down sampling of high frequency signal recordings to relatively low-resolution images, and the redundant information introduced by the presence of background pixels. These models had excellent performance on a validation dataset that was manually annotated and confirmed by cardiologists. The reported performance on the held-out test and external validation sets was limited by the quality of labels, which likely varied given higher performance on LBBB, RBBB, and AF, than for 1^st^ degree AV block, a pattern also observed in signal-based models.^3^ We confirmed this explicitly through expert ECG review of a sample of reportedly false positive ECGs – those where the model predicted a label with high confidence but the ECG was not actually labeled with the condition. We found that these were in fact true positives, representing incorrect labels.

The image-based model also learned to identify individual leads on varying ECG formats and identified lead-specific diagnostic cues, similar to human readers. In addition to external validation performance suggesting that the model did not train to rely on dataset-specific heuristics, the interpretability of the AI-based predictions further support their generalizability.^17^ Our examination of mean heatmaps across a sample of the most confident predictions for RBBB and LBBB showed that classifications for these intraventricular conduction disturbances were guided by the same information human readers focus on when reading an ECG. For example, diagnosis of RBBB in both forms of images was guided by the recognition of a widened S wave in lead V4, and an RSR’ pattern in leads V1 and V2. Similarly, diagnosis of LBBB in images was guided by focus on leads I, V3, and V4, where characteristic patterns emerge from monophasic R waves, QS complexes, and rS complexes, respectively. Similar heatmaps applied to individual ECGs further supported a similar interpretable learning across clinical labels.

Such interpretability addresses some of the implementation challenges of AI-based ECG models that are not readily explainable.^21^ Our findings also suggest that Grad-CAM can ameliorate this issue in real-time by providing an automated label, but also informing a clinician visually of the portions of the ECG that were used by the model to ascribe the label. Providing a Grad-CAM output in addition to the diagnosis and confidence of the diagnosis can provide context to predictions made by CNNs and aid in their acceptance in the clinical workflow.^1,22^

Our study has certain limitations. First, while our model has excellent performance characteristics, the reason for the discordance of the model and the labels could not be confirmed for all ECGs. The ECGs have been reviewed by a cardiologist in our training data,^3^ and by two cardiologists in the external validation data,^15^ but we found that high probability predictions initially noted to be false positives in both these sets actually represented inaccurate labels. Moreover, this pattern was not observed in the cardiologist-validated internal test set, further suggesting that the performance of the model likely exceeds what is reported in the held-out and external validation sets. Second, we focused on 6 clinical labels, based on their availability in the training data, but our models would not apply to other clinical disorders. To address this, follow up investigations will focus on developing models on repositories with broader sets of labels. Finally, we were unable to discern an interpretable pattern from Grad-CAM on ECG-based identification for gender classification, but the performance of the model on external validation data argues against overfitting. Moreover, Grad-CAM does not allow for interpretability of rhythm disorders, where large sections of repeating patterns in the ECG, which cannot be defined by heatmaps, are required for diagnosis. While the model performed well on these labels, they represent a challenge for the interpretability of AI applications on ECG tracings.

## CONCLUSIONS

We have developed an externally validated multilabel automated diagnosis algorithm that accurately identifies rhythm and conduction disorders from either ECG images or raw electrocardiographic signals. The versatility, interpretability, generalizability, and broad ability to incorporate ECG data of all formats has the potential to broaden the application of artificial intelligence to clinical electrocardiography.

## Data Availability

The data in the manuscript are available from the authors of the original study, Antonio H Ribeiro et al. Nat Communication. 2020 Apr 9;11(1):1760. The validation data are available from Physionet.org.

https://github.com/CarDS-Yale/ECG-DualNet

