## Supplementary material for "Automated Multilabel Diagnosis on Electrocardiographic Images and Signals": Online Supplement

### eMETHODS

#### Signal Model Parameters

Each convolution was padded so that input and output size was the same and was followed by a batch normalization layer and a ReLU activation layer. Inception blocks were composed of three parallel 1D convolutional blocks with kernels of size 40, 20, and 10, which were concatenated and followed by a pooling layer. The number of channels for the convolutional layers was block dependent and increased further into the model. Batch normalization was used to rescale signals after each convolutional layer, and dropout layers were added to prevent overfitting. The model was trained with an Adagrad optimizer and learning rate of $5 \times 10^{-3}$ for 10 epochs with a minibatch size of 64. Model weights were randomly initialized with Xavier initialization.

eTable 1: Dataset Characteristics

| | Cardiologist Validated Test Set | Held-Out Test Set | PTBXL | Training + Validation |
|---|---|---|---|---|
| n | 827 | 111412 | 21785 | 2116824 |
| Female, n (%) | 506 (61.2) | 67192 (60.3) | 10442 (47.9) | 1276674 (60.3) |
| age, mean (SD) | 54.9 (16.5) | 53.5 (17.5) | 59.8 (17.0) | 53.6 (17.4) |
| 1dAVb, n (%) | 28 (3.4) | 1722 (1.5) | 793 (3.6) | 32724 (1.5) |
| RBBB, n (%) | 34 (4.1) | 3031 (2.7) | 541 (2.5) | 58520 (2.8) |
| LBBB, n (%) | 30 (3.6) | 1734 (1.6) | 522 (2.4) | 32943 (1.6) |
| SB, n (%) | 16 (1.9) | 1808 (1.6) | 637 (2.9) | 33633 (1.6) |
| AF, n (%) | 13 (1.6) | 1983 (1.8) | 1507 (6.9) | 37679 (1.8) |
| ST, n (%) | 37 (4.5) | 2425 (2.2) | 825 (3.8) | 45871 (2.2) |

eTable 2: Class Balancing Weighting Scheme for Image and Signal-Based Models

| Label | Positive Class n (%) | Positive Class Weight | Negative Class Weight |
| --- | --- | --- | --- |
| Female | 1276674 (60.3) | 0.99456179 | 1.00543821 |
| 1dAVb | 32724 (1.5) | 1.72679131 | 0.27320869 |
| RBBB | 58520 (2.8) | 1.58113966 | 0.41886034 |
| LBBB | 32943 (1.6) | 1.72534378 | 0.27465622 |
| SB | 33633 (1.6) | 1.72058533 | 0.27941467 |
| AF | 37679 (1.8) | 1.69507613 | 0.30492387 |
| ST | 45871 (2.2) | 1.64671409 | 0.35328591 |

eTable 3: Performance of image models trained with and without shuffled images on standard and alternate format images in the held-out test set

| Training Scheme | Test Image Format | Label | Accuracy | PPV | NPV | Specificity | Sensitivity | AUROC | F1 | AUPRC |
|---|---|---|---|---|---|---|---|---|---|---|
| Shuffled | Standard | Male | 0.831 | 0.766 | 0.879 | 0.834 | 0.826 | 0.909 | 0.795 | 0.873 |
| | | 1dAVb | 0.98 | 0.39 | 0.992 | 0.988 | 0.486 | 0.968 | 0.433 | 0.372 |
| | | RBBB | 0.987 | 0.725 | 0.996 | 0.991 | 0.856 | 0.993 | 0.785 | 0.786 |
| | | LBBB | 0.993 | 0.734 | 0.997 | 0.995 | 0.832 | 0.996 | 0.78 | 0.805 |
| | | SB | 0.985 | 0.534 | 0.996 | 0.989 | 0.778 | 0.989 | 0.633 | 0.581 |
| | | AF | 0.989 | 0.668 | 0.996 | 0.993 | 0.755 | 0.988 | 0.709 | 0.737 |
| | | ST | 0.986 | 0.624 | 0.997 | 0.989 | 0.848 | 0.993 | 0.719 | 0.693 |
| | | Weighted Mean | 0.987 | 0.625 | 0.996 | 0.991 | 0.774 | 0.989 | 0.691 | 0.678 |
| | Alternate | Male | 0.835 | 0.767 | 0.887 | 0.833 | 0.839 | 0.913 | 0.802 | 0.877 |
| | | 1dAVb | 0.98 | 0.381 | 0.992 | 0.987 | 0.506 | 0.967 | 0.435 | 0.37 |
| | | RBBB | 0.988 | 0.752 | 0.995 | 0.992 | 0.826 | 0.992 | 0.788 | 0.796 |
| | | LBBB | 0.993 | 0.765 | 0.997 | 0.996 | 0.81 | 0.996 | 0.787 | 0.806 |
| | | SB | 0.985 | 0.534 | 0.997 | 0.989 | 0.796 | 0.99 | 0.639 | 0.588 |
| | | AF | 0.989 | 0.67 | 0.995 | 0.993 | 0.751 | 0.989 | 0.708 | 0.744 |
| | | ST | 0.987 | 0.646 | 0.996 | 0.99 | 0.842 | 0.994 | 0.731 | 0.714 |
| | | Weighted Mean | 0.987 | 0.639 | 0.995 | 0.991 | 0.768 | 0.989 | 0.695 | 0.686 |
| Unshuffled | Standard | Male | 0.83 | 0.762 | 0.881 | 0.83 | 0.83 | 0.908 | 0.795 | 0.871 |
| | | 1dAVb | 0.977 | 0.356 | 0.993 | 0.984 | 0.569 | 0.968 | 0.438 | 0.375 |
| | | RBBB | 0.988 | 0.738 | 0.996 | 0.992 | 0.843 | 0.992 | 0.787 | 0.785 |
| | | LBBB | 0.993 | 0.75 | 0.997 | 0.996 | 0.818 | 0.996 | 0.782 | 0.796 |
| | | SB | 0.985 | 0.527 | 0.996 | 0.988 | 0.784 | 0.988 | 0.63 | 0.561 |
| | | AF | 0.988 | 0.647 | 0.996 | 0.992 | 0.771 | 0.988 | 0.704 | 0.735 |
| | | ST | 0.986 | 0.623 | 0.997 | 0.988 | 0.856 | 0.993 | 0.721 | 0.71 |
| | | Weighted Mean | 0.986 | 0.622 | 0.996 | 0.99 | 0.785 | 0.988 | 0.691 | 0.677 |
| | Alternate | Male | 0.836 | 0.772 | 0.883 | 0.839 | 0.831 | 0.912 | 0.801 | 0.876 |

| | | | | | | | | | | |
|---|---|---|---|---|---|---|---|---|---|---|
| | | 1dAVb | 0.979 | 0.372 | 0.993 | 0.986 | 0.531 | 0.967 | 0.437 | 0.373 |
| | | RBBB | 0.987 | 0.722 | 0.996 | 0.991 | 0.868 | 0.992 | 0.789 | 0.795 |
| | | LBBB | 0.993 | 0.731 | 0.997 | 0.995 | 0.841 | 0.996 | 0.783 | 0.801 |
| | | SB | 0.985 | 0.522 | 0.997 | 0.988 | 0.815 | 0.989 | 0.636 | 0.593 |
| | | AF | 0.989 | 0.715 | 0.994 | 0.995 | 0.677 | 0.989 | 0.696 | 0.73 |
| | | ST | 0.986 | 0.627 | 0.997 | 0.988 | 0.869 | 0.994 | 0.729 | 0.725 |
| | | Weighted Mean | 0.987 | 0.628 | 0.996 | 0.99 | 0.781 | 0.989 | 0.693 | 0.686 |

eTable 4: Performance of image models trained with and without shuffled images on standard and alternate format images in the cardiologist-validated test set

| Training Scheme | Test Image Format | Label | Accuracy | PPV | NPV | Specificity | Sensitivity | AUROC | F1 | AUPRC |
|---|---|---|---|---|---|---|---|---|---|---|
| Shuffled | Standard | Male | 0.775 | 0.669 | 0.874 | 0.739 | 0.832 | 0.845 | 0.742 | 0.775 |
| | | 1dAVb | 0.985 | 0.808 | 0.991 | 0.994 | 0.75 | 0.989 | 0.778 | 0.744 |
| | | RBBB | 0.982 | 0.721 | 0.996 | 0.985 | 0.912 | 0.979 | 0.805 | 0.832 |
| | | LBBB | 0.996 | 0.966 | 0.997 | 0.999 | 0.933 | 1 | 0.949 | 0.989 |
| | | SB | 0.992 | 0.737 | 0.998 | 0.994 | 0.875 | 0.995 | 0.8 | 0.756 |
| | | AF | 0.992 | 0.75 | 0.995 | 0.996 | 0.692 | 0.996 | 0.72 | 0.786 |
| | | ST | 0.987 | 0.81 | 0.996 | 0.99 | 0.919 | 0.997 | 0.861 | 0.927 |
| | | Weighted Mean | 0.988 | 0.808 | 0.995 | 0.992 | 0.867 | 0.992 | 0.833 | 0.857 |
| | Alternate | Male | 0.771 | 0.656 | 0.893 | 0.711 | 0.866 | 0.859 | 0.746 | 0.786 |
| | | 1dAVb | 0.978 | 0.692 | 0.988 | 0.99 | 0.643 | 0.984 | 0.667 | 0.708 |
| | | RBBB | 0.987 | 0.829 | 0.994 | 0.992 | 0.853 | 0.987 | 0.841 | 0.858 |
| | | LBBB | 0.994 | 0.963 | 0.995 | 0.999 | 0.867 | 0.999 | 0.912 | 0.971 |
| | | SB | 0.993 | 0.857 | 0.995 | 0.998 | 0.75 | 0.995 | 0.8 | 0.73 |
| | | AF | 0.994 | 0.786 | 0.998 | 0.996 | 0.846 | 0.998 | 0.815 | 0.89 |
| | | ST | 0.994 | 0.921 | 0.997 | 0.996 | 0.946 | 0.996 | 0.933 | 0.906 |
| | | Weighted Mean | 0.99 | 0.851 | 0.994 | 0.995 | 0.829 | 0.993 | 0.839 | 0.854 |
| Unshuffled | Standard | Male | 0.779 | 0.705 | 0.829 | 0.804 | 0.738 | 0.842 | 0.721 | 0.77 |
| | | 1dAVb | 0.976 | 0.605 | 0.994 | 0.981 | 0.821 | 0.981 | 0.697 | 0.709 |
| | | RBBB | 0.984 | 0.839 | 0.99 | 0.994 | 0.765 | 0.987 | 0.8 | 0.857 |
| | | LBBB | 0.996 | 0.935 | 0.999 | 0.997 | 0.967 | 0.999 | 0.951 | 0.983 |
| | | SB | 0.989 | 0.684 | 0.996 | 0.993 | 0.812 | 0.995 | 0.743 | 0.765 |
| | | AF | 0.992 | 1 | 0.991 | 1 | 0.462 | 0.992 | 0.632 | 0.725 |
| | | ST | 0.985 | 0.857 | 0.991 | 0.994 | 0.811 | 0.996 | 0.833 | 0.924 |

| | | | | | | | | | | |
|---|---|---|---|---|---|---|---|---|---|---|
| | | Weighted Mean | 0.986 | 0.818 | 0.993 | 0.993 | 0.804 | 0.992 | 0.799 | 0.85 |
| | **Alternate** | Male | 0.807 | 0.714 | 0.884 | 0.787 | 0.838 | 0.874 | 0.771 | 0.812 |
| | | 1dAVb | 0.977 | 0.622 | 0.994 | 0.982 | 0.821 | 0.983 | 0.708 | 0.706 |
| | | RBBB | 0.989 | 0.838 | 0.996 | 0.992 | 0.912 | 0.997 | 0.873 | 0.935 |
| | | LBBB | 0.995 | 0.964 | 0.996 | 0.999 | 0.9 | 0.999 | 0.931 | 0.974 |
| | | SB | 0.992 | 0.765 | 0.996 | 0.995 | 0.812 | 0.996 | 0.788 | 0.763 |
| | | AF | 0.993 | 0.769 | 0.996 | 0.996 | 0.769 | 0.996 | 0.769 | 0.829 |
| | | ST | 0.992 | 0.917 | 0.995 | 0.996 | 0.892 | 0.997 | 0.904 | 0.92 |
| | | Weighted Mean | 0.989 | 0.829 | 0.995 | 0.993 | 0.867 | 0.995 | 0.845 | 0.872 |

eTable 5: Performance of signal models with and without peak morphology inputs on the held-out test set

| Model | Label | Accuracy | PPV | NPV | Specificity | Sensitivity | AUROC | F1 | AUPRC |
|---|---|---|---|---|---|---|---|---|---|
| **Signal + Peak Morphology** | Male | 0.704 | 0.594 | 0.829 | 0.641 | 0.8 | 0.803 | 0.682 | 0.737 |
| | 1dAVb | 0.968 | 0.214 | 0.991 | 0.976 | 0.411 | 0.934 | 0.282 | 0.197 |
| | RBBB | 0.987 | 0.733 | 0.995 | 0.992 | 0.827 | 0.987 | 0.777 | 0.745 |
| | LBBB | 0.992 | 0.697 | 0.997 | 0.994 | 0.821 | 0.994 | 0.754 | 0.754 |
| | SB | 0.986 | 0.561 | 0.996 | 0.99 | 0.767 | 0.99 | 0.648 | 0.589 |
| | AF | 0.984 | 0.535 | 0.994 | 0.99 | 0.643 | 0.979 | 0.584 | 0.567 |
| | ST | 0.986 | 0.639 | 0.997 | 0.989 | 0.854 | 0.992 | 0.731 | 0.708 |
| | Weighted Mean | 0.984 | 0.584 | 0.995 | 0.989 | 0.738 | 0.981 | 0.649 | 0.615 |
| **Signal** | Male | 0.716 | 0.612 | 0.824 | 0.674 | 0.781 | 0.808 | 0.686 | 0.743 |
| | 1dAVb | 0.97 | 0.238 | 0.991 | 0.979 | 0.427 | 0.939 | 0.306 | 0.224 |
| | RBBB | 0.987 | 0.722 | 0.996 | 0.991 | 0.853 | 0.988 | 0.782 | 0.757 |
| | LBBB | 0.992 | 0.696 | 0.997 | 0.994 | 0.837 | 0.994 | 0.76 | 0.764 |
| | SB | 0.987 | 0.571 | 0.996 | 0.991 | 0.762 | 0.991 | 0.653 | 0.599 |
| | AF | 0.985 | 0.585 | 0.993 | 0.992 | 0.625 | 0.98 | 0.604 | 0.603 |
| | ST | 0.986 | 0.638 | 0.997 | 0.989 | 0.858 | 0.993 | 0.732 | 0.71 |
| | Weighted Mean | 0.985 | 0.594 | 0.995 | 0.99 | 0.745 | 0.982 | 0.659 | 0.63 |

eTable 6: Performance of signal models with and without peak morphology inputs on the cardiologist-validated test set

| Model | Label | Accuracy | PPV | NPV | Specificity | Sensitivity | AUROC | F1 | AUPRC |
|---|---|---|---|---|---|---|---|---|---|
| **Signal + Peak Morphology** | Male | 0.712 | 0.594 | 0.847 | 0.646 | 0.816 | 0.795 | 0.688 | 0.703 |
| | 1dAVb | 0.97 | 0.538 | 0.991 | 0.977 | 0.75 | 0.97 | 0.627 | 0.607 |
| | RBBB | 0.99 | 0.842 | 0.997 | 0.992 | 0.941 | 0.994 | 0.889 | 0.881 |
| | LBBB | 0.996 | 0.966 | 0.997 | 0.999 | 0.933 | 0.997 | 0.949 | 0.967 |
| | SB | 0.989 | 0.667 | 0.998 | 0.991 | 0.875 | 0.996 | 0.757 | 0.814 |
| | AF | 0.99 | 0.667 | 0.996 | 0.994 | 0.769 | 0.986 | 0.714 | 0.746 |
| | ST | 0.994 | 0.944 | 0.996 | 0.997 | 0.919 | 0.998 | 0.932 | 0.952 |
| | Weighted Mean | 0.988 | 0.803 | 0.996 | 0.992 | 0.88 | 0.991 | 0.836 | 0.848 |
| **Signal** | Male | 0.713 | 0.597 | 0.841 | 0.656 | 0.804 | 0.803 | 0.685 | 0.711 |
| | 1dAVb | 0.971 | 0.577 | 0.984 | 0.986 | 0.536 | 0.96 | 0.556 | 0.501 |
| | RBBB | 0.99 | 0.882 | 0.995 | 0.995 | 0.882 | 0.997 | 0.882 | 0.928 |
| | LBBB | 0.995 | 1 | 0.995 | 1 | 0.867 | 0.998 | 0.929 | 0.97 |
| | SB | 0.989 | 0.667 | 0.998 | 0.991 | 0.875 | 0.995 | 0.757 | 0.686 |
| | AF | 0.994 | 1 | 0.994 | 1 | 0.615 | 0.993 | 0.762 | 0.802 |
| | ST | 0.993 | 0.878 | 0.999 | 0.994 | 0.973 | 0.998 | 0.923 | 0.923 |
| | Weighted Mean | 0.989 | 0.837 | 0.994 | 0.994 | 0.816 | 0.99 | 0.82 | 0.824 |

eFigure 1: Efficientnet-B3 Architecture used for Image Model

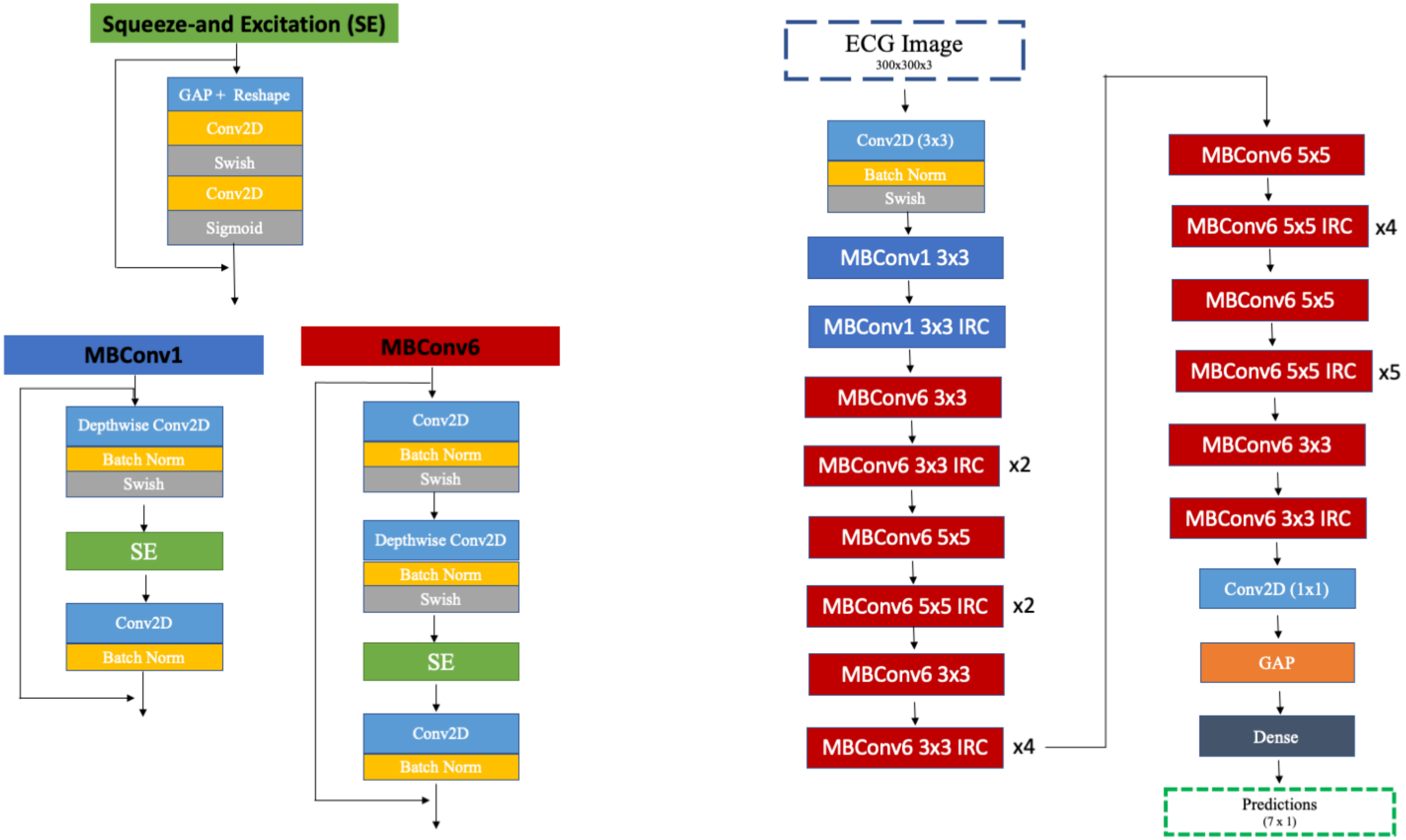

eFigure 2: Custom Inception Architecture used for Signal Model

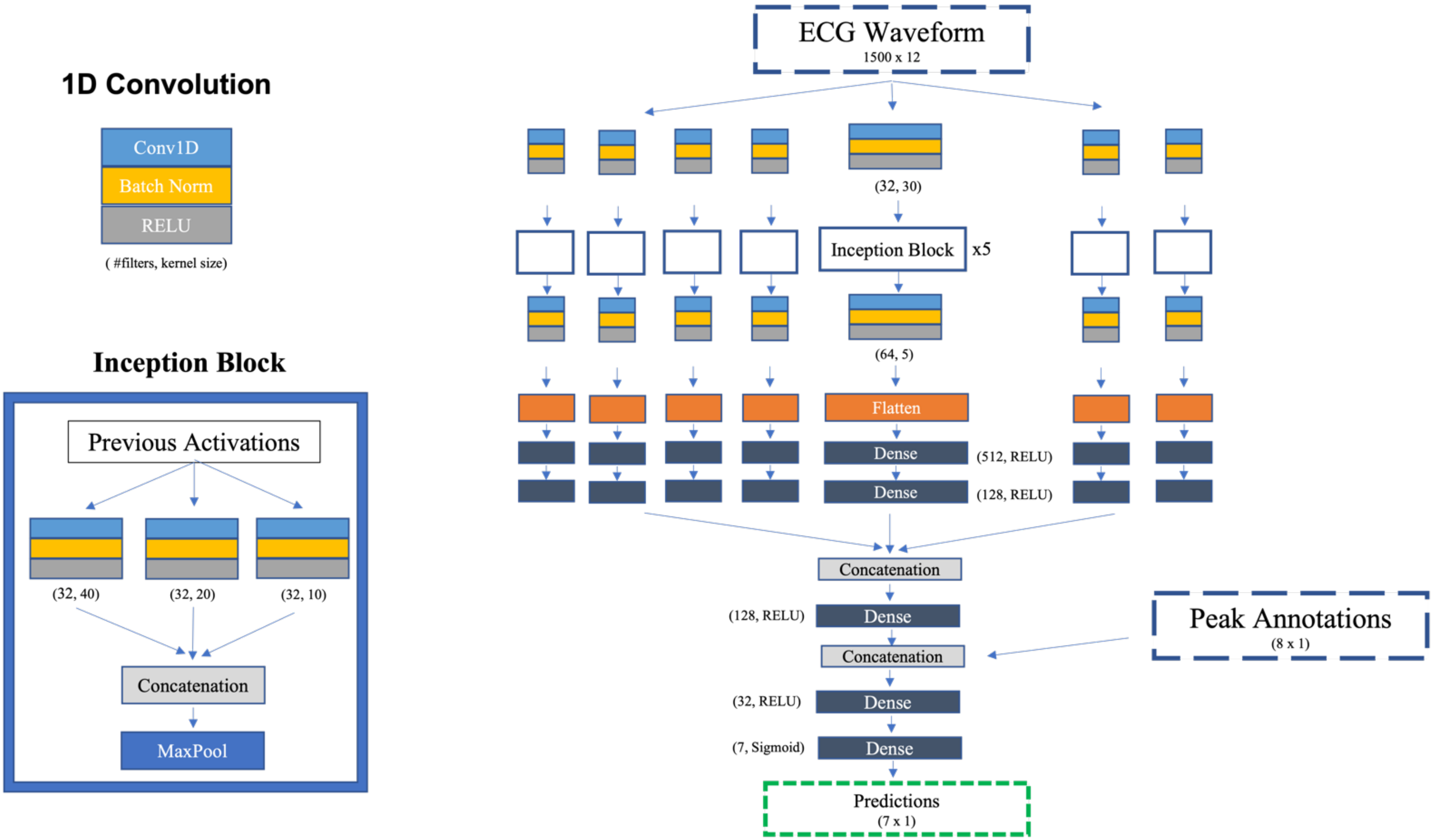

eFigure 3: Confusion Matrices for Signal Model Predictions

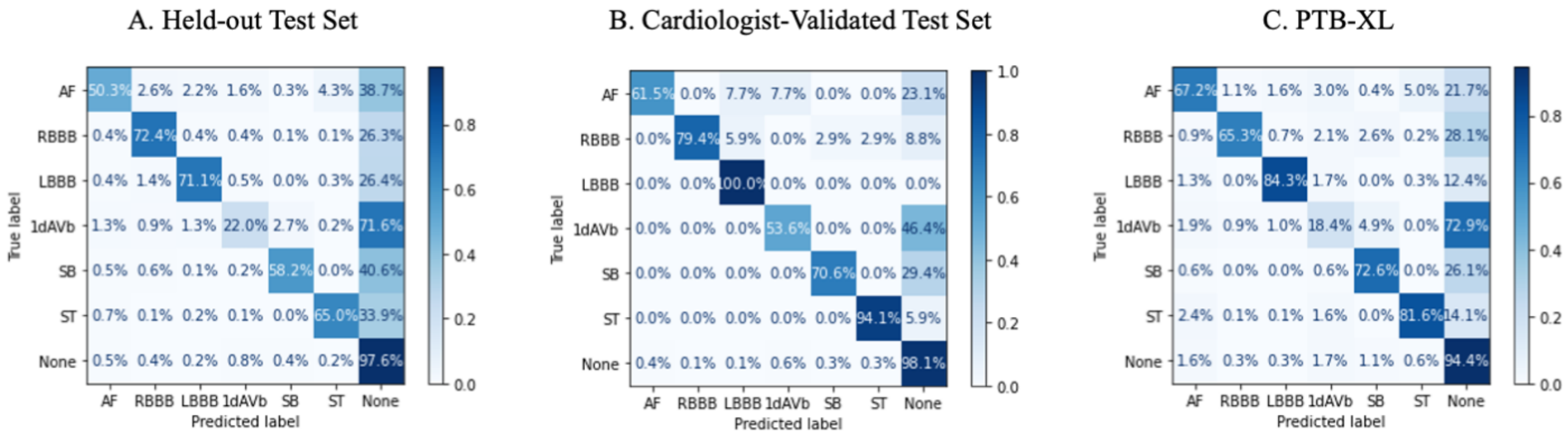

eFigure 4: Representative Grad-CAMs

A. RBBB

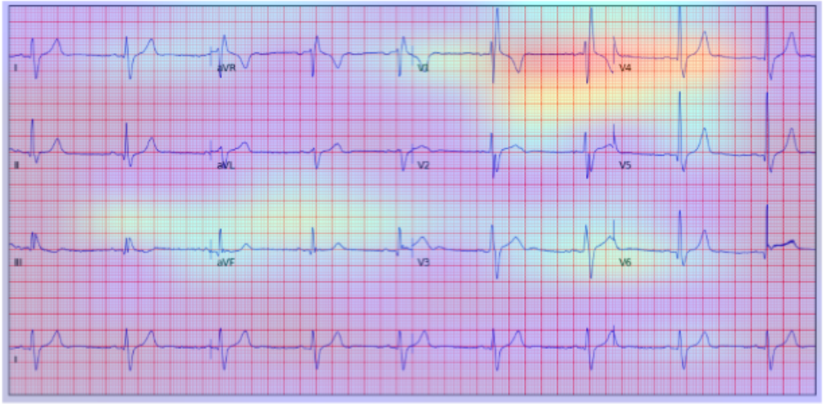

B. LBBB

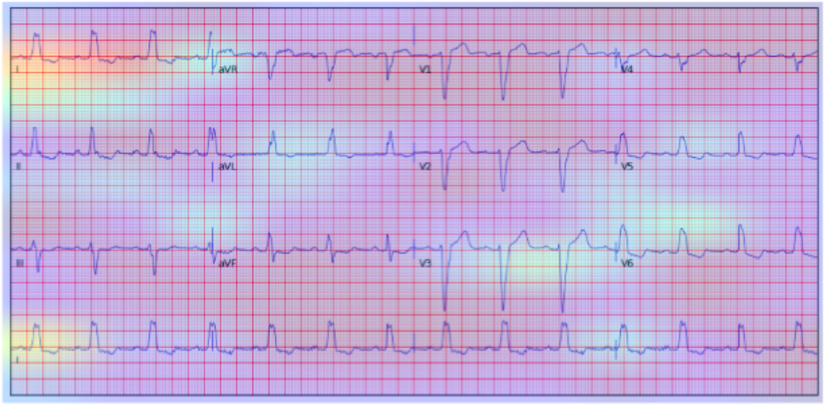

C. Sinus Bradycardia

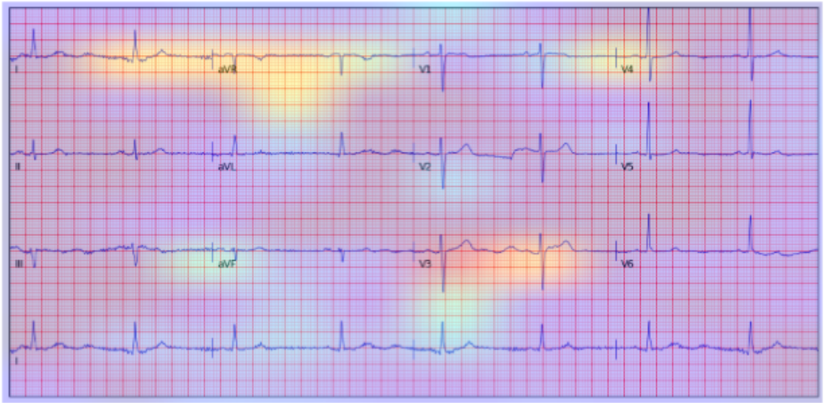

D. Sinus Tachycardia

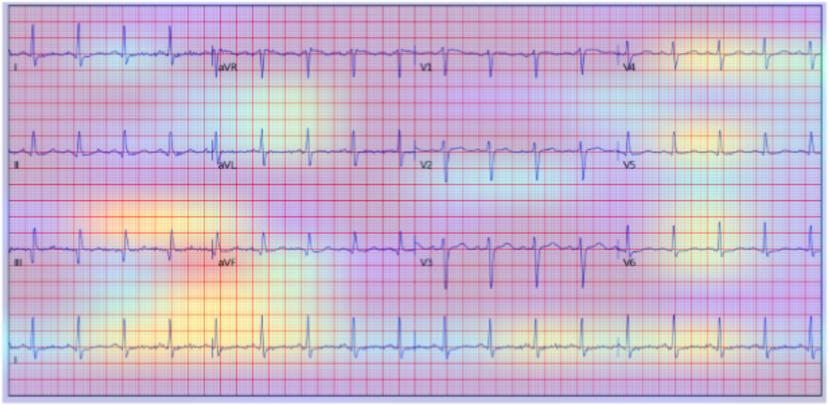
